# Safety of Tenecteplase in Pediatric Arterial Ischemic Stroke: Results from an International Surveillance Study

**DOI:** 10.64898/2026.07.20.26358531

**Authors:** Sarah Lee, Lisa R. Sun, Jacqueline Lee-Eng, Dwight Barry, Jenny L Wilson, Dana B Harrar, Marcela Torres, Maria M. Galardi, Sahar M Hassanein, Megan Barry, Michael J Rivkin, Kristin Guilliams, Catherine Amlie-Lefond, the International Pediatric Stroke Study and the Pediatric Neurocritical Care Research Group

## Abstract

**Background:** Recent AHA guidelines recommend the use of IV Tenecteplase (TNK) in adult stroke patients, but there is minimal safety and dosing data on TNK in pediatric patients. We performed a safety surveillance study aiming to evaluate risk of symptomatic intracranial hemorrhage (ICH) in children who received TNK for suspected acute ischemic stroke (AIS).

**Methods:** This was a prospective observational cohort surveillance study analyzing responses from a monthly email survey sent to members of two large international pediatric stroke and neurocritical care research consortia querying recent use of TNK in children. Limited demographic, clinical, and outcome data were collected. A Bayesian beta-binomial model for risk of symptomatic ICH with IV TNK was fit using a prior distribution based on the risk level in adults.

**Results:** Between February 2023-June 2026, 44 children received TNK for suspected AIS. Most patients (n=37, 84.1%) were adolescents; no children under 5 received TNK. Twelve patients (27.3%) were ultimately diagnosed with a stroke mimic. Symptomatic ICH was not reported in any children who received IV TNK; 3 had asymptomatic ICH on follow-up imaging. One patient received intra-arterial TNK and experienced symptomatic ICH. No other major bleeding events were reported.

**Conclusions:** IV TNK is being administered to pediatric patients with suspected stroke in clinical practice, and may be safe in older children with AIS. Rigorous prospective studies are needed to better assess risk and outcomes in this unique population.

## Introduction

Intravenous (IV) thrombolysis is standard of care in adults with acute arterial ischemic stroke (AIS). The 2026 American Heart Association/American Stroke Association (AHA/ASA) Guideline for the Early Management of Patients With Acute Ischemic Stroke equally recommends IV tenecteplase (TNK) or alteplase for adult patients presenting in early and extended windows, based on the strength of multiple large randomized controlled trials demonstrating noninferiority of TNK’s efficacy and safety compared to alteplase.^1,2^ TNK has the added benefits of better fibrin specificity and more efficient administration in a single 5-10 second bolus compared to alteplase, which requires a bolus followed by 1-hour infusion. Many hospitals worldwide are replacing alteplase with TNK in their adult stroke protocols based on mounting evidence and recommendations, but there is persistent uncertainty on how best to treat children.

For the first time, the 2026 AHA/ASA guidelines specifically address pediatric patients, stating that IV alteplase may be considered in children aged 28 days to 18 years with confirmed AIS “as it is safe, but efficacy is uncertain.”^3^ The dearth of safety and dosing data for IV TNK is discussed in the text of the pediatric thrombolysis section, and no recommendations are made. However, while some stroke centers have opted to continue using IV alteplase for pediatric cases citing more robust safety data, others have followed the lead of their integrated or affiliated adult stroke centers and transitioned to TNK as the single thrombolytic agent for patients of all ages— including children—with the goal of streamlining protocols, simplifying pharmacy requests and reducing potential dosing and administration errors. Furthermore, children with stroke sometimes present to adult emergency departments and receive the thrombolytic available in that facility. Thus, children are receiving and will continue to receive IV TNK as part of clinical practice for acute stroke. Given the known differences in hemostasis and stroke pathophysiology between children and adults, safety and dosing data are urgently needed.

The aim of this study is to report updated demographic, clinical and safety data from a prospective, international surveillance study on the use of TNK for suspected AIS in children. The primary safety outcome of interest was symptomatic intracranial hemorrhage (sICH).

## Methods

Since December 2023, monthly email notifications with a link to an online prospective TNK surveillance survey have been sent to members of the International Pediatric Stroke Study (IPSS) and the Pediatric Neurocritical Care Research Group (PNCRG), two large research consortia comprised of medical providers dedicated to children with stroke and neurocritical illness. Together, IPSS and PNCRG have over 100 participating centers across 34 countries. Survey recipients are encouraged to forward the link to interested colleagues. The study is ongoing.

Study approval was obtained through the Institutional Review Board at Seattle Children’s Hospital, with a waiver of consent due to the limited dataset and lack of patient identifying information. The survey queries whether the completing provider has cared for a child who received TNK in the preceding month, and gathers limited information on patient demographics, outcomes, and complications. All responses are collected in a secure REDCap database. The full survey and detailed methods have been reported previously.^4^ The complete list of data elements collected in the survey is included in Appendix 1.

### Statistical Analysis

Descriptive statistics were used to report overall survey results, comparison between stroke and stroke mimics, and comparison with existing literature on TNK. A Bayesian beta-binomial model was used to assess the probability of risk of sICH following IV TNK in children. The prior was modeled as a beta (1.5, 16.63) distribution, which has a mode of 3.1%, the estimated risk in adults. To test for robustness, the model was also analyzed with a uniform prior of beta (1, 1), which implies that all values of risk could be equally likely. Risk estimates are reported as posterior medians and 95% quantile credible intervals (CrI).^1^ All statistical analyses were performed using R, version 4.4 (R Core Team [2025]; R: A Language and Environment for Statistical Computing; R Foundation for Statistical Computing, Vienna, Austria).

Anonymized data not published herein will be made available by reasonable request from any qualified investigator.

## Results

Between February 2023 and June 2026, 730 total survey submissions were received, and 52 surveys reported patients who received TNK. Of those, 5 were duplicate entries and 3 were 18 years of age or older, leaving 44 unique pediatric patients who received TNK included in our analysis. Over half (n=26, 59.1%) were female. The majority (n=37, 84.1%) were adolescents between 13-17 years of age and 7 patients (15.9%) were between 5-12 years of age; no patients under 5 years of age were reported. Twelve patients (27.3%) were ultimately diagnosed with a stroke mimic. Comparison between children with confirmed stroke versus stroke mimic are shown in Table 1. Most survey respondents were neurologists (n=37), followed by intensivists (n=5) and hematologists (n=2). Nine children were reported to have received TNK in 2023, 14 in 2024, 17 in 2025, and 4 in 2026 (January-June). Enrollment over the study period is shown in Figure 1.

**Figure 1.**
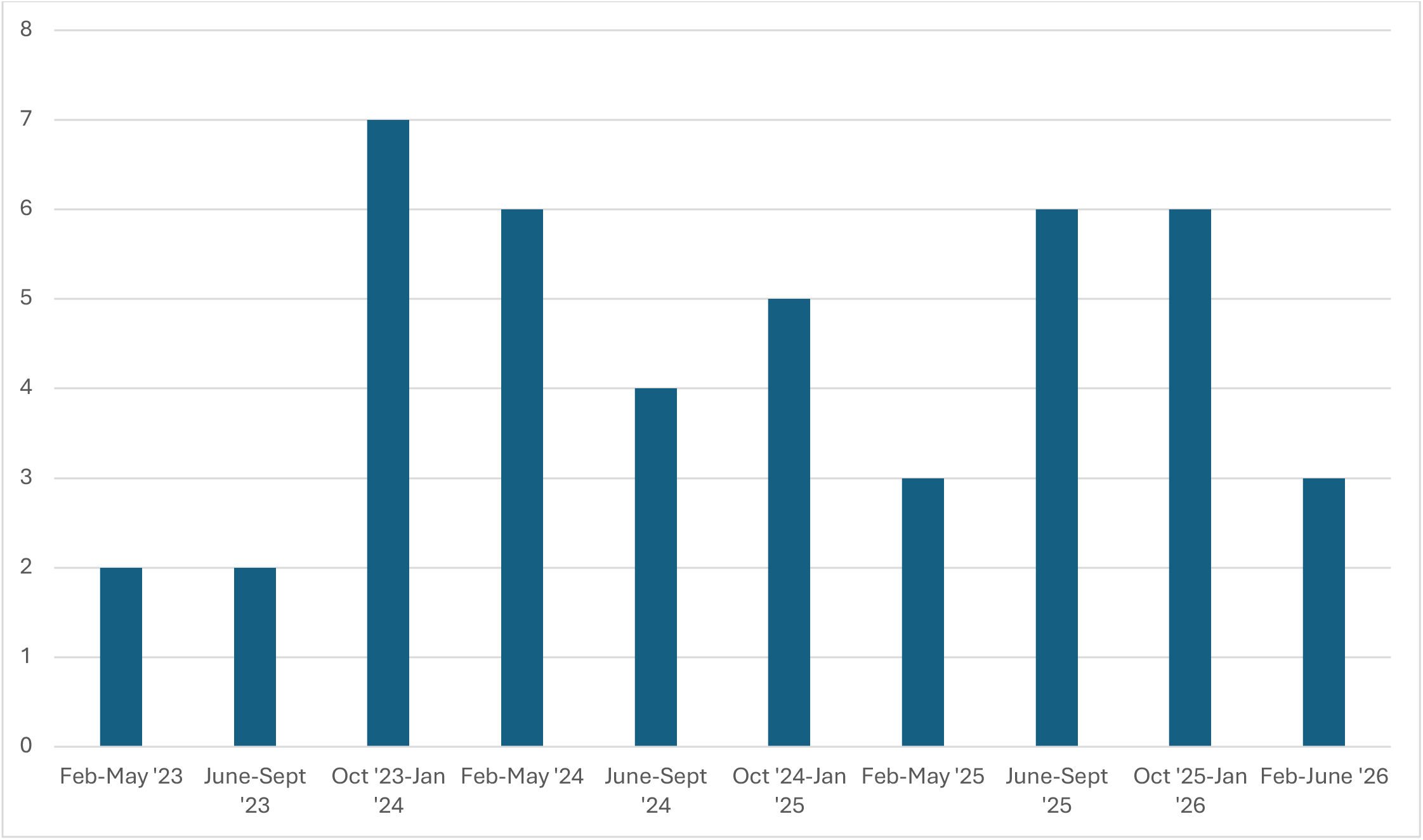
Number of children reported to have received TNK (y-axis) over the study period (x-axis).

**Table 1.** Characteristics of children with stroke versus stroke mimic in children who received TNK.

|  | All patients<br>n=44 | Stroke<br>n=32 | Stroke mimic<br>n=12 |
| --- | --- | --- | --- |
| Female, n (%) | 26 (59.1) | 17 (53.1) | 9 (75) |
| <5 years, n (%) | 0 (0) | 0 (0) | 0 (0) |
| 5-12 years, n (%) | 7 (15.9) | 5 (15.6) | 2 (16.7) |
| 13-17 years, n (%) | 37 (84.1) | 27 (84.4) | 10 (83.3) |
| Time from last known well or<br>symptom onset |  |  |  |
| 0-3 hours | 25 (56.8) | 19 (59.4) | 6 (50.0) |
| 3-4.5 hours | 18 (41.0) | 12 (37.5) | 6 (50.0) |
| >6 hours | 1 (2.3) | 1 (3.1) | 0 (0) |
| Location of TNK administration |  |  |  |
| Enrolling institution | 7 (15.9) | 6 (18.8) | 1 (8.3) |
| Outside institution | 37 (84.1) | 26 (81.2) | 11 (91.7) |
| Asymptomatic ICH | 3 (6.8) | 3 (9.4) | 0 (0) |
| Symptomatic ICH | 1* | 1* | 0 (0) |
| Discharge Location |  |  |  |
| Home | 33 (75.0) | 23 (71.9) | 10 (83.3) |
| Rehabilitation | 11 (25.0) | 9 (28.1) | 2 (16.7) |

\*This patient received IA TNK at a dose of 0.11 mg/kg

Most patients (n=37, 84.1%) received TNK at an external facility prior to transfer to the reporting institution; 29/37 (78.4%) were transferred from an adult hospital, 1/37 (2.7%) was transferred from a pediatric hospital, and the remaining 7 did not specify the type of institution the patient was transferred from. All 8 of the sites who gave TNK at their institution were pediatric hospitals. TNK was given between 0-3 hours for 25 patients (56.8%); between 3-4.5 hours in 18 patients (40.9%) and beyond 6 hours in 1 patient (2.3%). Of the 29 adult hospitals that gave TNK, 17/29 (58.6%) did so within 0-3 hours and the remainder between 3-4.5 hours; of the 9 pediatric centers that gave TNK, 4 (44.4%) did so within 0-3 hours and the remainder (65.6%) between 3-4.5 hours.

Forty-three patients received IV TNK; of those, 42/43 were given the same dosage as adults (0.25mg/kg, max 25mg). One patient was given a 50mg IV dose for treatment of concurrent pulmonary embolism and stroke, and one patient was reported to have received intra-arterial (IA) TNK at a dose of 0.11mg/kg.

Nearly all patients (43/44, 97.7%) underwent follow-up imaging. Of those, one patient had sICH and 3 patients had asymptomatic intracranial hemorrhage (aICH) on follow-up imaging; all intracranial hemorrhages were in children with confirmed stroke. The sICH was reported in the patient who received IA TNK and was in the 5-12-year-old age group. The 3 aICH patients had received IV TNK at the 0.25mg/kg dose; 2 were 13-17 years old and 1 was 5-12 years old. The estimated risk of sICH for IV TNK in children based on zero incidents in 43 patients is 2.0% (95% CrI 0.2-7.5%), using a prior assumption of 3.1% risk from adults, or 1.6% (95% CrI 0.1-8.0%) based on no prior assumptions about risk (Figure 2).

**Figure 2.**
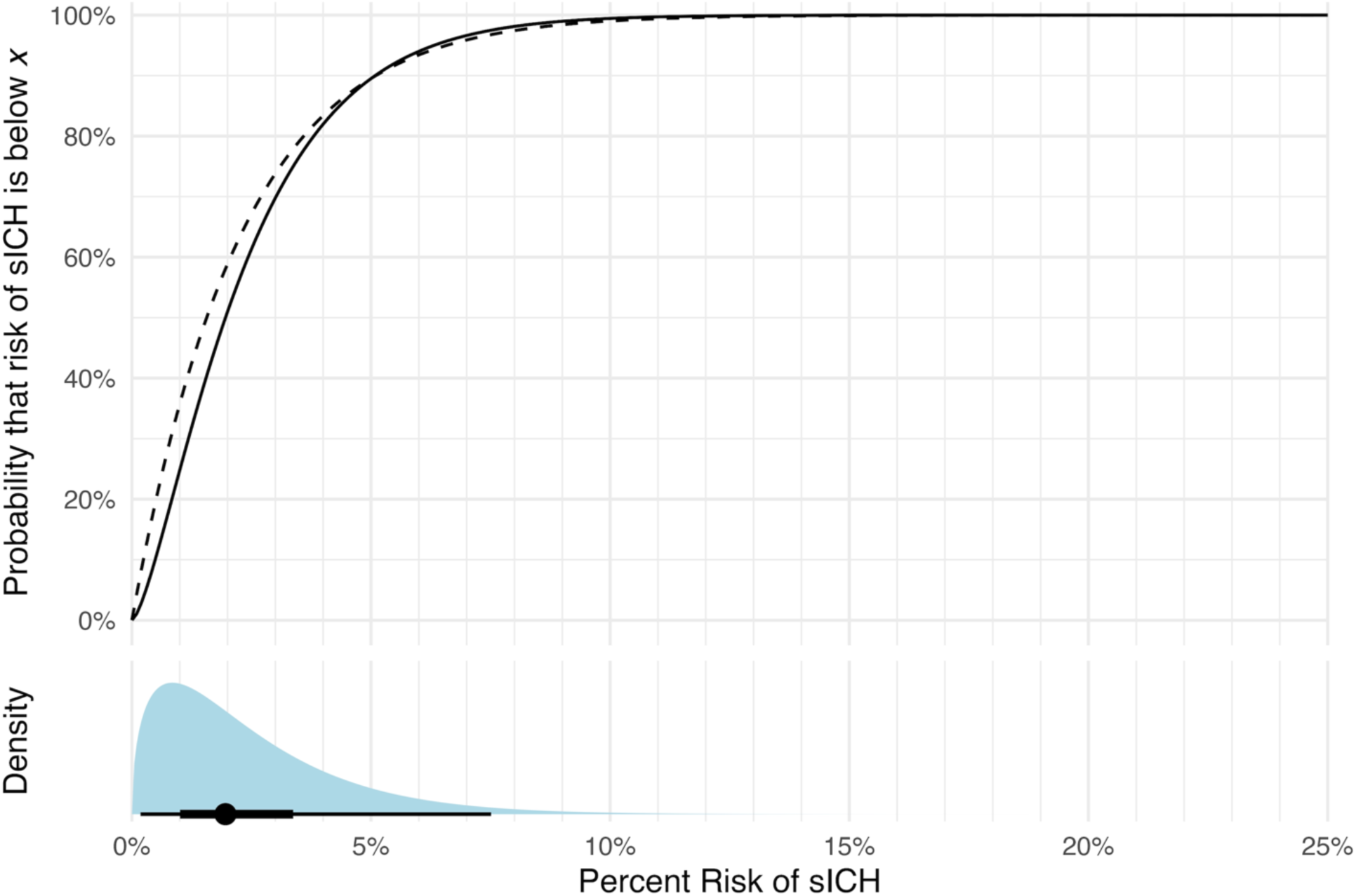
Estimated probability of sICH after IV TNK based on a Bayesian beta-binomial regression of zero sICH incidents in 44 patients with a 3.1% adult risk prior (solid curve, upper panel) and a uniform prior (dashed curve, upper panel). To interpret the curves, read up from x axis to the curve and then to the y axis to determine the probability (y) that the true risk is at or below (x). Lower panel shows the probability density for the 3.1% risk prior and the median (point), 50% credible interval (thick line), and 95% credible interval (thin line) of the risk estimate.

Presumed stroke etiology was reported in 14/32 patients (43.8%) with confirmed stroke, and included cardioembolism (n=5), focal cerebral arteriopathy (n=2), paradoxical embolism (n=2), multifactorial etiology (n=2), cryptogenic (n=2), and primary central nervous system vasculitis (n=1). Discharge destination was home for 33/44 patients (75%) and to a rehabilitation facility for 11/44 (25%). Among patients who were discharged to rehab, 2/11 (18.1%) had a final diagnosis of stroke mimic.

## Discussion

Our data suggest that IV TNK may be safe in older children when given at the standard adult dosage of 0.25mg/kg. Importantly, no children under age 5 were reported to have received IV TNK in this cohort; most cases (>80%) were 13 years of age or older, with only ∼16% in the 5-12-year-old age group. Furthermore, given the broad age range categories, it is not known whether all or most of the children in the 5-12-year-old age group were closer to 12 years of age versus 5 years of age; thus, the safety of IV TNK in younger children cannot be inferred. In the TIPSTERS study investigating the use of IV alteplase in children, no pediatric patients younger than 1 year of age received thrombolysis, emphasizing the lack of hyperacute treatment data in the infant and toddler age group.^5^ Other published case series on IV alteplase in children similarly have minimal to no data on children under 2 years (Table 2).^6–8^ The explanation for the age discrepancy is likely multifactorial: first, providers may feel more reluctant to administer an off-label treatment to younger children, whose stroke etiologies and general cerebrovascular and hemodynamic physiology differ significant from adolescents and adults; second, younger children may bypass an adult hospital (where the majority of TNK is being given) and instead go directly to a pediatric facility, where IV TNK may not be part of the treatment protocol; and third, determining treatment eligibility may be challenging for younger children, for whom precise time of symptom onset and magnitude of clinical deficit is often difficult to assess.

**Table 2.** Existing studies of thrombolysis with IV alteplase children.

| Author | Type of<br>study | # patients<br>treated with IV<br>alteplase | # patients<br><1 year of<br>age | # patients<br>between 1-3 years of<br>age | Youngest<br>patient<br>reported<br>(years) |
| --- | --- | --- | --- | --- | --- |
| Amlie-Lefond et<br>al, *Stroke*,<br>2020.<sup>5</sup> | Retrospective | 26 | 0 | 1 | 1 |
| Kossorotoff et<br>al, JAMA<br>Network, 2022.<sup>6</sup> | Retrospective | 44 total (28 IV<br>alteplase alone,<br>16 IV alteplase<br>prior to EVT) | 0 | 0 | 4 |
| Bigi et al, Ann<br>Neurol, 2018<sup>7</sup> | Retrospective | 5 | 0 | 0 | 5 |
| Tabone et<br>al, *Stroke*, 2017<sup>8</sup> | Retrospective | 11 | 0 | 0 | 3 |

EVT= endovascular thrombectomy

Nearly all patients in our study received IV TNK within 4.5 hours, and over half within 3 hours of last known well, indicating that children are presenting and being treated within a hyperacute timeframe. Earlier pediatric stroke studies reported that diagnosis of pediatric stroke is often delayed due to lack of awareness and difficulty obtaining emergent imaging, among other factors.^9^ More recent studies, however, suggest that many pediatric patients—particularly those with large vessel occlusion—present within a timeframe that qualifies them for treatment with IV thrombolysis or endovascular thrombectomy (EVT).^10^ Additionally, most patients in our series were treated with TNK at an outside facility prior to transfer. Notably, over half of the adult hospitals and none of the pediatric hospitals administered TNK within the earliest time window of 0-3 hours. This may be because adult hospitals see a higher volume of stroke patients and have streamlined systems to provide treatment more rapidly. These data may be helpful when considering feasibility for pediatric treatment trials that might require screening within a narrow time window.

Interestingly, one patient in our cohort received IV TNK >6 hours after last known well. Extended window thrombolysis (>4.5 hours) with either IV TNK or alteplase is now considered for certain adult patients with evidence of salvageable tissue on perfusion imaging or DWI-FLAIR mismatch on MRI. Children have more robust pial collaterals than older adults and thus theoretically may have an even longer time window for treatment; however, it is not known whether the same mismatch eligibility criteria that have been investigated in adults are applicable to children.^3,11^ Giving thrombolysis to a patient with an established stroke may increase the risk of sICH. In our cohort, the patient who received TNK beyond 6 hours did not report sICH, aICH, or other major bleeding concerns, though further study is required to evaluate the utility of IV thrombolysis, and under what conditions, in children in extended time windows.

The one patient who received IA TNK was the only patient reported to have sICH. Due to the limited nature of data collection, circumstances surrounding this patient’s presentation and treatment (i.e. magnitude of infarct burden, clinical deficit, and/or whether they also underwent EVT) are not known. The current adult guidelines state that, for adult patients with AIS who undergo successful EVT (defined as a modified Thrombolysis in Cerebral Infarction score ≥2b), adjunctive IA TNK “may be reasonable to improve cerebral perfusion and 90-day functional outcome.” That said, results from clinical trials are mixed, and the role of IA TNK in adult AIS is controversial.^12–14^ Of note, the dose of IA TNK used in adult trials was 0.0625mg/kg, lower than the reported dose administered to the pediatric patient in our study (0.11mg/kg). More research is needed to evaluate safety, dosing, and efficacy of IA TNK in younger patients.

Stroke mimics are more common in children presenting with focal neurologic deficits compared with adults.^15,16^ Over 25% of children who received IV TNK in our study were diagnosed with a stroke mimic—higher than most adult estimates, which range from 1-16%.^17,18^ Importantly, none of the children with stroke mimic experienced intracranial hemorrhage, consistent with adult data on IV thrombolysis, which indicates that the hemorrhage rate in stroke mimics is low—roughly 0.5-1.2%, compared to 2-8% in patients with stroke.^19,20^ Because neither IV alteplase nor TNK is FDA-approved for children and because stroke mimics are highly prevalent, evidence that the event is a true stroke is often preferable in children before administering off-label treatment with limited safety data. The AHA/ASA guidelines specifically state that IV alteplase may be considered in pediatric patients “with confirmed AIS,” in contrast to adult recommendations where the potential benefit of faster treatment for a patient with disabling deficits outweighs the risk of treating a stroke mimic. Thus, many pediatric stroke protocols in North America prefer MRI as the initial imaging modality over CT—both for its superior sensitivity for acute ischemia as well as the lack of exposure to ionizing radiation.^21^ Understanding the implications of treating stroke mimics in children is an area that requires further study.

Our study has several important limitations. To maximize the number and timeliness of responses, we significantly limited the level of detail in the survey questions to maintain anonymity. As a result, we were unable to gather more granular information regarding patient demographics, eligibility criteria, and/or clinical, laboratory, imaging, and outcome data that might enhance our understanding of TNK this patient population. The survey was circulated monthly to members of two large research consortia with an international membership; thus, its respondents were more likely to be specialized providers invested in pediatric stroke and neurocritical care, potentially influencing outcomes and not necessarily generalizable to under-resourced or non-academic settings. Survey participation was voluntary and thus prone to reporting and severity bias; furthermore, longer-term adverse effects and patient clinical outcome data were not solicited. Finally, as previously mentioned, there were few patients younger than 12 years and no patients younger than 5 years reported, thus safety results cannot be extrapolated to these age groups.

Nevertheless, this study represents an important step in understanding the use of IV TNK in children. We found that children—mostly adolescents—<u>are</u> receiving TNK for acute AIS as well as for stroke mimics, with a favorable safety profile overall. Rigorous prospective studies are still urgently needed to understand optimal dosing, safety, and efficacy of TNK in pediatric patients with AIS.

## Acknowledgements

We thank our colleagues for their participation in the online prospective Tenecteplase in childhood stroke surveillance survey.

## Appendix 1: Information collected in the tenecteplase (TNK) in Pediatrics Survey

Name and affiliation of individual completing the survey

Role of submitting provider in the patient’s care: Neurologist, Intensivist, Neurosurgeon, Radiologist, Neurosurgeon, Nurse, Other

Whether TNK was given in previous month for stroke/suspected stroke.

If TNK was given:

Whether it was given in their institution or outside hospital prior to transport.
Type of outside hospital where given: Adult hospital, Pediatric hospital

Patient sex: Male, Female, Other/nonbinary

Age range of the patient: < 1 month, 1 month – 2 years, 2 years – 5 years, 5 years – 12 years, 13 years – 17 years, 18 years +

Time from patient’s last seen well to TNK administration: 0-3 hours, 3-4.5 hours, 4.5-6 hours, >6 hours

Does of TNK administered: 0.25 mg/kg, 0.4 mg/kg, Other (free text)

Any major safety concern attributable to the TNK administration: Allergic reaction, Symptomatic intracranial hemorrhage, Other life-threatening hemorrhage, Systemic hemorrhage, Other safety concern, None.

Presence of intracranial bleeding on follow-up imaging: Symptomatic intracranial hemorrhage, Asymptomatic intracranial hemorrhage, No intracranial bleeding on follow-up imaging, No follow-up imaging obtained

Site of discharge: Home, Rehabilitation, Death

Final diagnosis: Stroke, Mimic

